# Role of CRISPR-Cas in Modulating Efflux Pump Gene Expression in *Acinetobacter baumannii* Isolates from Clinical Samples

**DOI:** 10.1101/2025.08.23.25334291

**Authors:** Alya A. Rahi, Batool Kereem Mohammed

## Abstract

The CRISPR-Cas system serves as an adaptive immune defence in bacteria, protecting against foreign genetic elements. In *Acinetobacter baumannii*, efflux pumps are major contributors to multidrug resistance (MDR). This study investigates the potential regulatory role of the CRISPR-Cas system on efflux pump genes, specifically *adeB*, and its association with antibiotic resistance.

**Methods:** A total of 100 clinical specimens were collected from patients admitted to the Wound Unit at Al-Hilla Teaching Hospital between March and May 2025. Standard bacteriological methods were used for isolation and identification. Antimicrobial susceptibility testing (AST) was conducted using the disk diffusion technique and interpreted according to the Clinical and Laboratory Standards Institute (CLSI) 2025 guidelines. PCR assays were used to detect the presence of CRISPR-Cas system components and the *blaOXA-51* gene. Quantitative real-time PCR (qRT-PCR) was employed to assess the expression levels of the *adeB* efflux pump gene.

**Results:** Out of the 100 clinical samples (44 females and 55 males, aged 10–55 years), 15 (15%) isolates were confirmed as *A. baumannii*. AST results indicated high resistance rates to oxacillin (100%), benzylpenicillin (93.3%), erythromycin (73.3%), and tetracycline (66.7%). The isolates exhibited the highest sensitivity to tigecycline (93.3%), trimethoprim/sulfamethoxazole (93.3%), and rifampicin (86.7%). All isolates were positive for the *blaOXA-51* gene.Molecular analysis revealed that the I-Fb subtype of the *cas1* gene was present in 86.7% of the isolates. Expression profiling showed that *adeB* was overexpressed in 66.6% of the isolates. Notably, isolates harboring complete CRISPR-Cas components exhibited downregulation of *adeB*, suggesting a possible repressive regulatory effect of CRISPR-Cas on efflux pump expression.

**Conclusion:** This study demonstrates variability in the distribution of CRISPR-Cas elements among clinical *A. baumannii* isolates and suggests a potential inverse correlation between CRISPR-Cas system presence—particularly the I-Fb-cas1 subtype—and *adeB* efflux pump gene expression. These findings highlight the potential of CRISPR-Cas systems to modulate resistance mechanisms in *A. baumannii*, warranting further investigation into their therapeutic implications.

## Introduction

*Acinetobacter baumannii* is a Gram-negative opportunistic pathogen renowned for its remarkable ability to persist in both natural and healthcare environments. This resilience is attributed to its minimal nutritional requirements and exceptional environmental adaptability (Pompilio et al., 2021). It has emerged as a major cause of nosocomial infections, particularly in critically ill patients, due to its tendency to develop multidrug resistance (MDR). A key mechanism driving this resistance is the active expulsion of antibiotics via efflux pump systems, especially those encoded by the *adeABC* operon. Within this operon, the *adeB* gene encodes a crucial transporter protein belonging to the resistance-nodulation-division (RND) family of efflux pumps, playing a central role in mediating antibiotic resistance (Ahuatzin-Flores *et al*., 2024).Concurrently, bacteria have evolved an adaptive immune mechanism known as the Clustered Regularly Interspaced Short Palindromic Repeats and CRISPR-associated proteins (CRISPR-Cas) system. This system provides defense against foreign genetic elements such as plasmids and bacteriophages. Beyond its canonical role in adaptive immunity, recent studies suggest that the CRISPR-Cas system may also regulate bacterial gene expression, influence virulence, and impact antimicrobial resistance (Baqar *et al*., 2024). Notably, specific CRISPR-Cas subtypes have been hypothesized to interfere with horizontal gene transfer and modulate the expression of endogenous genes, including those associated with drug resistance, such as efflux pump components (Karah *et al*., 2021).

## 2. Methodology

### 2.1. Specimen Collection and Culturing

In this study, 15 clinical specimens were obtained from the Wound Unit out of a total of 100 collected samples. All specimens were acquired from patients (44 females and 55 males), aged between 10 and 55 years, who exhibited various wound-related symptoms. The samples were collected at Hilla Teaching Hospital during the period from March to May 2025. Immediately following collection, specimens were transported to the laboratory for the isolation and identification of *Acinetobacter baumannii*. The samples were cultured on Hi-Chromagar and incubated at 37°C for 18–24 hours.

### 2.2. Antimicrobial Susceptibility Testing by Disk Diffusion Technique

Antimicrobial susceptibility testing (AST) was performed using the disk diffusion technique with 13 antibiotic discs supplied by Bioanalyse (Turkey). The antibiotics tested included:

- Benzylpenicillin (25 µg),Oxacillin (30 µg),Gentamicin (30 µg),Ciprofloxacin (30 µg)
- Erythromycin (10 µg),Rifampicin (10 µg),Imipenem (10 µg),Tetracycline (30 µg),Vancomycin (5 µg),Clindamycin (10 µg),Trimethoprim/Sulfamethoxazole (10 µg)
- Linezolid (10 µg),Tigecycline (30 µg).The diameters of inhibition zones were measured and interpreted according to the Clinical and Laboratory Standards Institute (CLSI) guidelines (2025).

### 2.3. DNA Extraction and Conventional PCR for CRISPR-Cas and *bla_OXA-51_* Gene Detection

Genomic DNA was extracted using a commercial kit (Geneaid Co., Taiwan), following the manufacturer’s protocol. The conventional polymerase chain reaction (PCR) was conducted to detect CRISPR-Cas subtype genes (I-Fa-cas1, I-Fb-cas1, I-Fa-cas3, I-Fb-cas3) and the *blaOXA-51* resistance gene. PCR amplification conditions included an initial denaturation at 94°C for 5 minutes, followed by 30 cycles of denaturation at 94°C for 1 minute, annealing for 30 seconds at gene-specific temperatures (see Table 1), and extension at 72°C for 1 minute. A final extension step was performed at 72°C for 10 minutes. Primer sequences and expected product sizes are listed in Table 1.

**Table (1):**
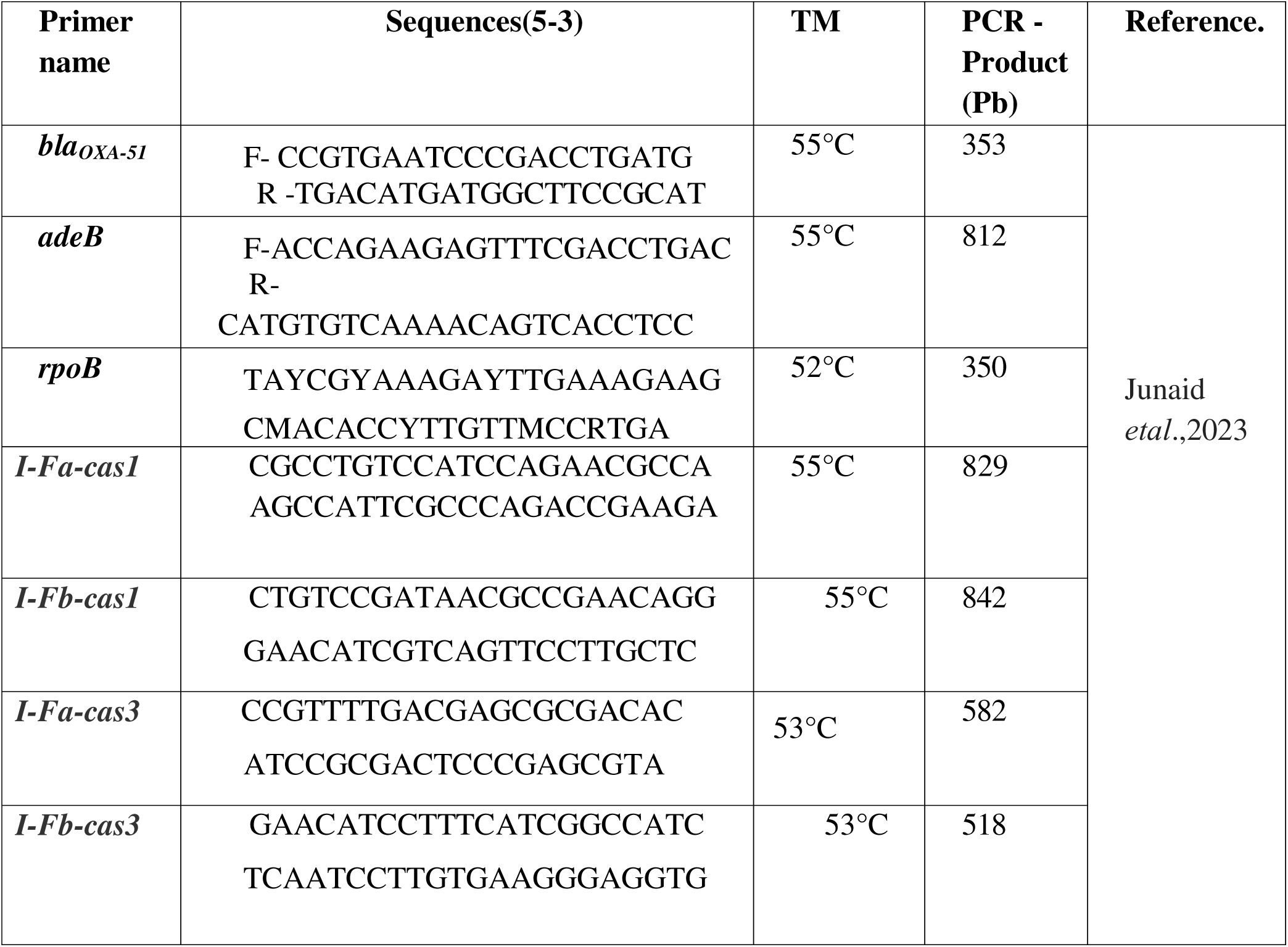
The primers and the sequences, Amplcon, TM, used in this study.

### 2.4. RNA Extraction and cDNA Synthesis

Total RNA was extracted using standard protocols such as TRIzol reagent or column-based purification kits, ensuring phase separation, effective washing, and integrity preservation. RNA concentration and purity were evaluated using spectrophotometric methods. High-quality RNA was then used for complementary DNA (cDNA) synthesis, employing reverse transcription with oligo(dT) primers. Proper reaction setup, incubation, and post-synthesis cleanup ensured the generation of reliable cDNA, which served as the template for downstream gene expression analyses.

### 2.5. Detection of CRISPR-Cas systems

PCR primers were used in this study, listed in Table (1).

#### Gene Expression of Efflux Pump Gene (*adeB*) by Quantitative RT-PCR

The q RT-PCR were calculated using the 2^^-ΔΔCt^ method.

#### CRISPR-Cas and Expression Efflux Pump Genes

#### Presence of CRISPR-Cas Loci and Its Potential Role in Antimicrobial Resistance

The presence of active CRISPR-Cas loci in clinical *Acinetobacter baumannii* isolates may suggest a reduced propensity for antimicrobial resistance (AMR). AMR in *A. baumannii* is commonly associated with the overexpression of multidrug efflux pumps—particularly those in the resistance-nodulation-division (RND) family, such as AdeABC, AdeIJK, and AdeFGH. These efflux systems contribute to resistance by actively expelling antibiotics from the bacterial cell. While the CRISPR-Cas system is classically recognised for its role in adaptive immunity against foreign genetic elements such as plasmids and bacteriophages, emerging evidence indicates its involvement in endogenous gene regulation, including genes linked to antimicrobial resistance. This suggests that CRISPR-Cas components may modulate the expression of efflux pump genes, potentially serving as intrinsic regulators of resistance mechanisms. Such findings open avenues for future research into the CRISPR-Cas system as a target for mitigating drug resistance in *A. baumannii*.

#### Statistical Analysis

All statistical analyses were performed using IBM SPSS Statistics, version 25.0. The significance level was set at *P* < 0.05. The data were evaluated using the independent samples *t*-test, and results with *P*-values below this threshold were considered statistically significant.

#### Inclusion Criteria

1. Clinical specimens obtained from patients with confirmed bacterial infections (e.g., bacteremia, urinary tract infection, pneumonia).
2. Samples collected specifically from wound swabs.
3. Inclusion of patients across all age groups and genders.
4. Patients who had not received antibiotic therapy for a minimum of 72 hours prior to sample collection.
5. Samples that adhered to standard protocols for collection and transport to prevent contamination.

#### Exclusion Criteria

1. Samples obtained from patients who had received antibiotics within three days prior to collection.
2. Nonclinical or improperly collected specimens (e.g., contaminated or of insufficient volume).
3. Samples from patients with infections caused by organisms other than *A. baumannii*.
4. Duplicate or repeat samples from the same patient within a short timeframe, unless justified by clinical necessity.
5. Specimens lacking complete clinical or demographic data.

#### Patient Consent Statement

All participants were informed of the study’s objectives and procedures. Written informed consent was obtained from each patient or their legal guardian prior to inclusion in the study.

#### .Ethical approval

Ethical approval for this study was obtained from the ethical committee at Hilla Surgical Teaching Hospital. This study was also approved by a local ethics committee at the College of Medicine, University of Babylon. and the hospital ethics committee under document number [ [IRB: 399-4/4/2025].

## 3. Results

### 3.1. Isolation and identification of *A.baumannii* isolates

A total of 15(15%) out of 100 clinical specimens of wound swabs from the hospitals in Babylon (Hilla Teaching Hospital) during the period from March 2025 to May 2025. while 30% were other bacterial sp. And 55% negative culture as shown in Figure 1.

**Figure (1):**
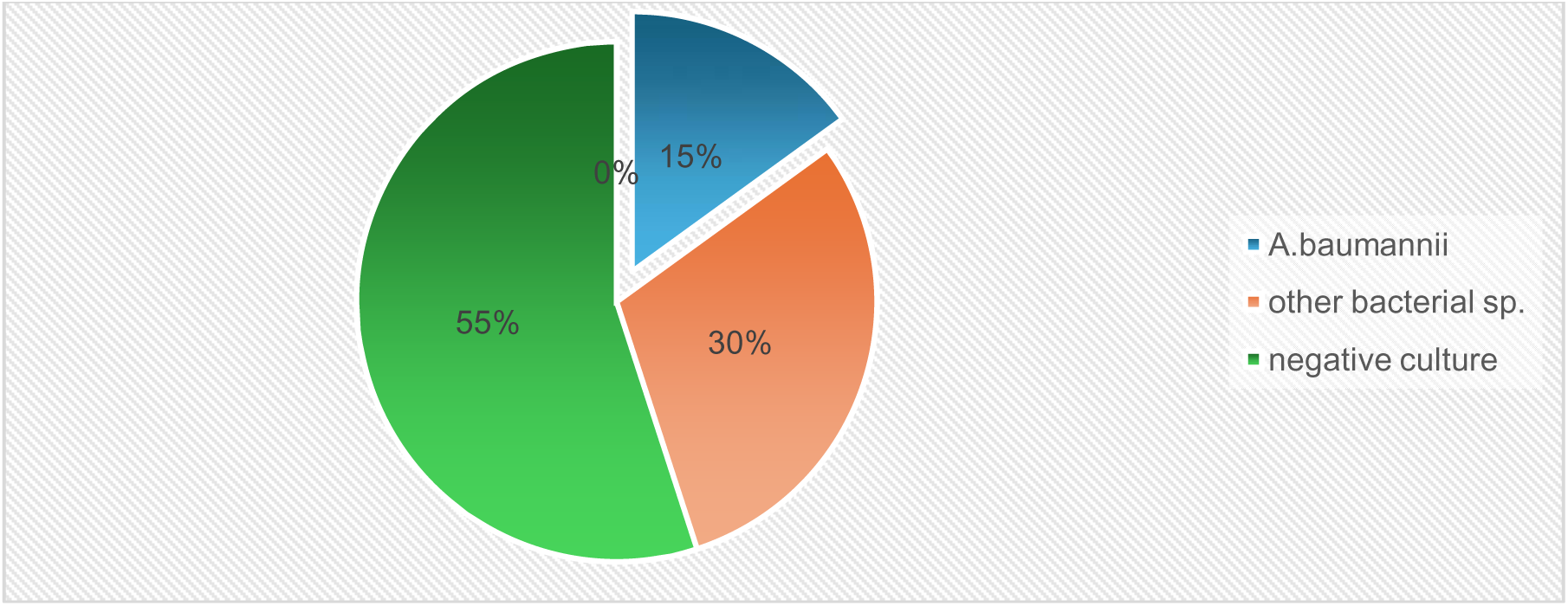
Percentage of Isolation rate in *A.baumannii* from wound swabs of clinical specimens

### 3.2. Antibiotic Susceptibility Testing

In this study, 15 *Acinetobacter baumannii* isolates were cultured on Mueller-Hinton agar and tested for susceptibility against 13 antibiotics. The findings revealed the following: High Resistance Rates: The isolates demonstrated notably high resistance to several antibiotics, including benzylpenicillin (93.3%), oxacillin (100%), erythromycin (73.3%), and tetracycline (66.7%). Resistance was also observed for ciprofloxacin (53.3%) and clindamycin (60%). High Sensitivity Rates: Tigecycline and trimethoprim/sulfamethoxazole both exhibited excellent activity, with sensitivity rates of 93.3%. Rifampicin also showed strong efficacy at 86.7%. Additional effective antibiotics included linezolid (86.7%), vancomycin (80%), and teicoplanin (73.7%). Intermediate Susceptibility: Some isolates demonstrated intermediate susceptibility to ciprofloxacin (20%) and vancomycin (13.3%), suggesting variability in response and potential treatment challenges.

**Table (2):**
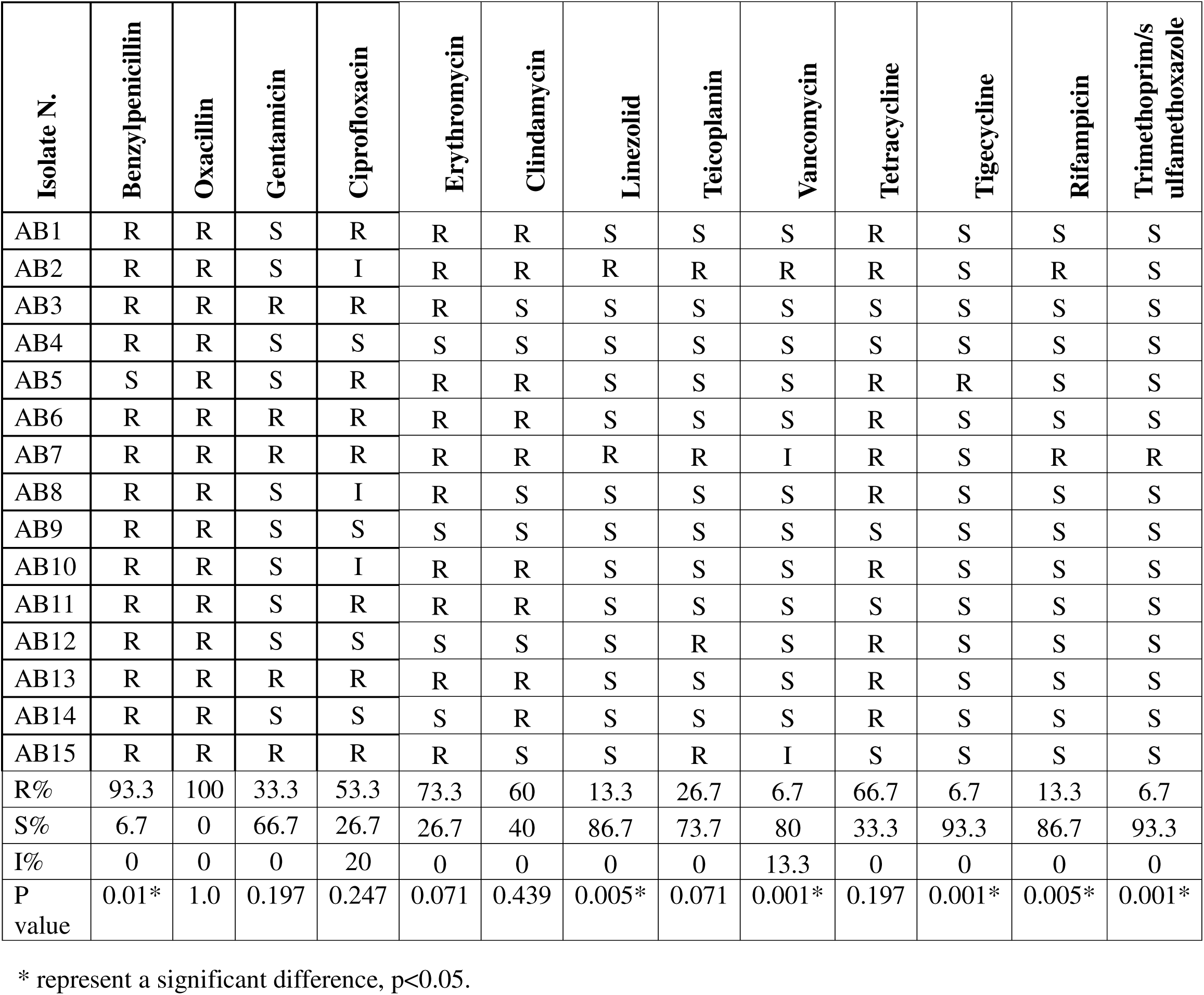
Antibiotic distribution percentages of 15 *Acinetobacter baumannii* isolates.

**Fig. (2):**
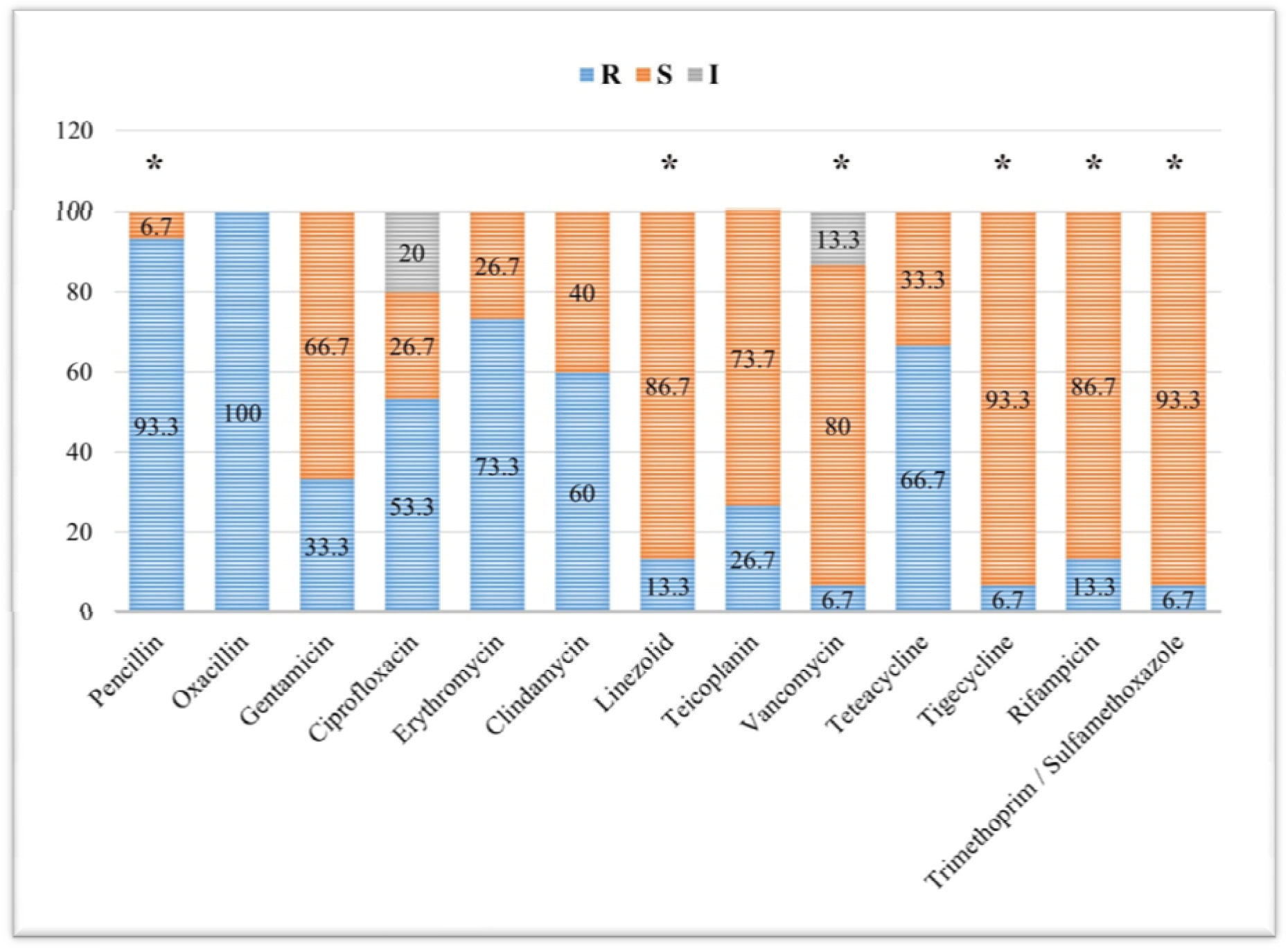
Antibiotic distribution percentages of 15 *Acinetobacter baumannii* clinical isolates. * represents a significant difference at p<0.05. R, resistant; S, sensitive; I, intermediate

### 3.3. Molecular detection of CRSIPR-Cas genes (*I-Fa-cas1*,*I-Fb-cas1*,*I-Fa-cas3,I-Fb-as3,blaOxa-51*) in *A.baumannii* isolates by polymerase chain reaction technique

In the present study, all 15 isolates (100%) tested positive for the *blaOXA-51* gene, as shown in Table 1. The *blaOXA-51* gene is intrinsic to *Acinetobacter baumannii* and serves as a reliable molecular marker for its identification. These results confirm the accurate classification of all isolates as *A. baumannii*.Regarding CRISPR-Cas system-associated genes:

> The I-Fa-cas1 gene was detected in 5 out of 15 isolates (33.3%) (Table 3). Its presence may be strain-specific or influenced by environmental or genomic factors.The I-Fb-cas1 gene was identified in 13 out of 15 isolates (86.67%) (Table 4). Its high prevalence and statistically significant association (Table 5) suggest that it could play an important role in *A. baumannii* and may serve as a potential target for future investigations into CRISPR-Cas functionality in this species.The I-Fa-cas3 gene was found in 6 of 15 isolates (40%) (Table 6), indicating moderate distribution and suggesting a possibly limited or context-specific function.The I-Fb-cas3 gene was detected in 8 of 15 isolates (53.33%) (Table 7). Although present in over half of the isolates, its distribution appears random, and no clear association with pathogenic traits was observed.These findings highlight the varying prevalence of CRISPR-Cas-related genes in *A. baumannii* and underscore the potential relevance of I-Fb subtype elements in this pathogen’s genomic landscape.

**Table (3):** Identification of *Acinetobacter baumannii* among isolates using *blaOXA51*-specific primer.

| Results | N | Percentage |
| --- | --- | --- |
| Positive | 15 | (100%) |
| Negative | 0 | (0.00%) |
| Total | 15 | 100% |

**Table (4):**
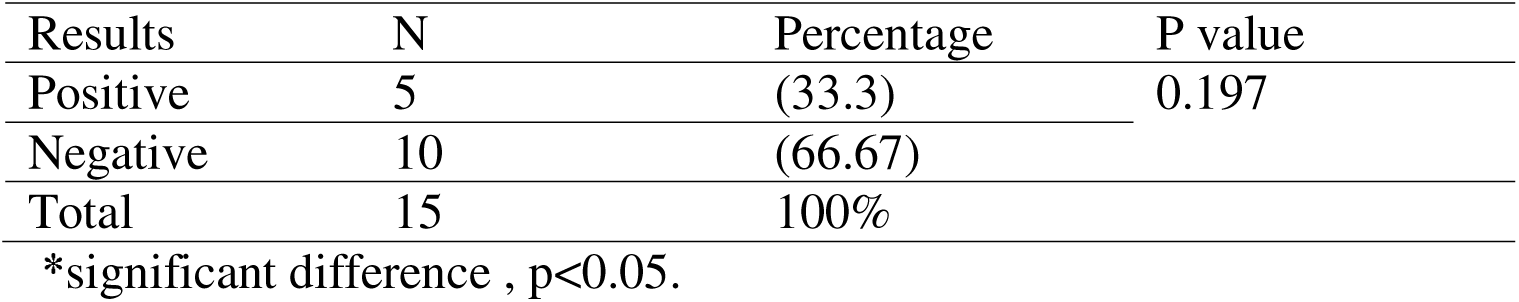
Identification of I-Fa-*cas1* gene among pathogenic *Acinetobacter baumannii*.

**Table (5):**
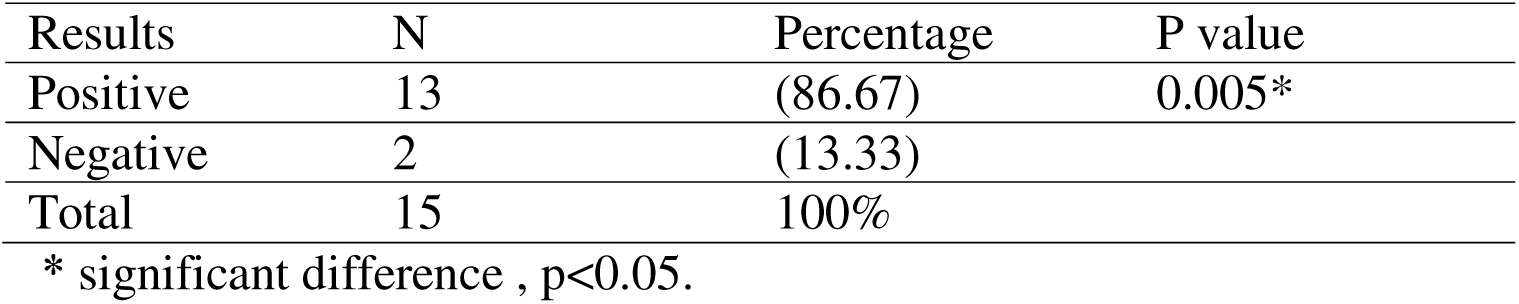
Identification of I-Fb-*cas1* gene among pathogenic *Acinetobacter baumannii* isolates.

**Table (6):** Identification of I-Fa-*cas3* gene among pathogenic *Acinetobacter baumannii*.

| Results | N | Percentage | P value |
| --- | --- | --- | --- |
| Positive | 6 | (40) | 0.439 |
| Negative | 9 | (60) |  |
| Total | 15 | 100% |  |

\*significant difference , $p < 0.05$ .

**Table (7):**
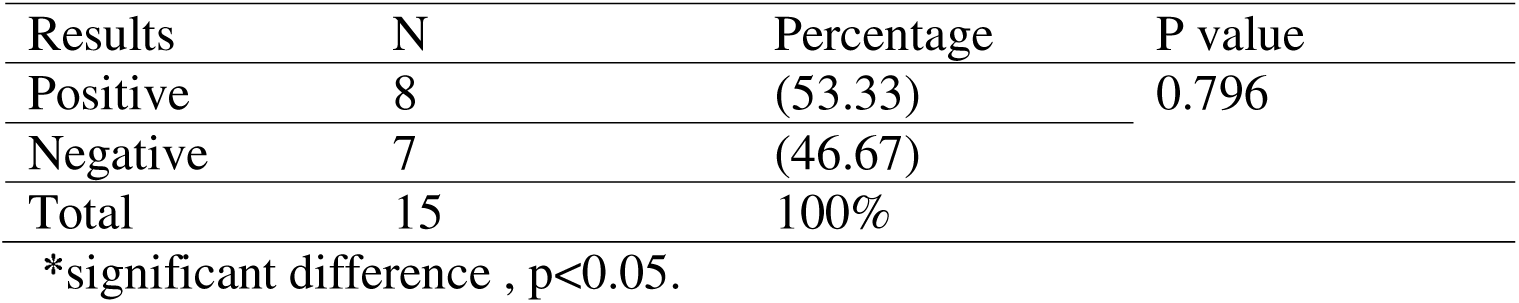
Identification of I-Fb-*cas3* gene among pathogenic *Acinetobacter baumannii*.

**Fig (3):**
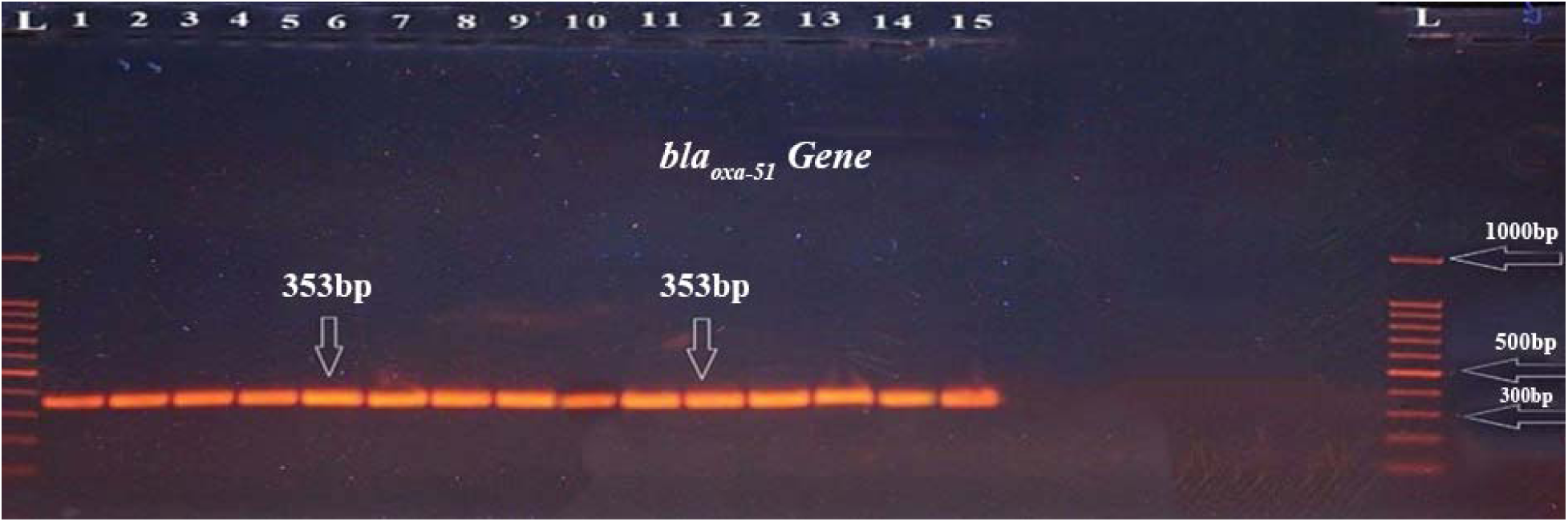
Gel electrophoresis of PCR product of *bla_OXA-51_*(353 bp) gene,L = ladder (100 -1500), melting temperature (Tm) of 55°C, 1% agarose gel, starting at 100 volts for 10 minutes and then reduced to 70 volts for 60 minutes, the 1–25 represents of the positive results in samples for this gene.

**Fig (4):**
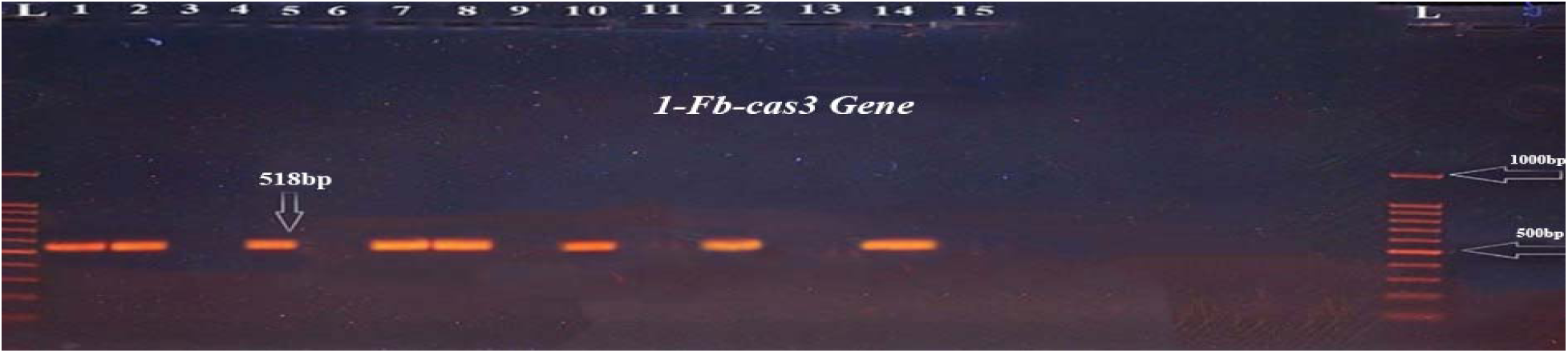
Gel electrophoresis of PCR product of *1-Fb-cas3*(518 bp) gene,L = ladder (100 -1500), melting temperature (Tm) of 55°C, 1% agarose gel, starting at 100 volts for 10 minutes and then reduced to 70 volts for 60 minutes, the (1,2,5,7,8,10,12,14 isolates represents of the positive results in samples for this gene.

**Fig (5):**
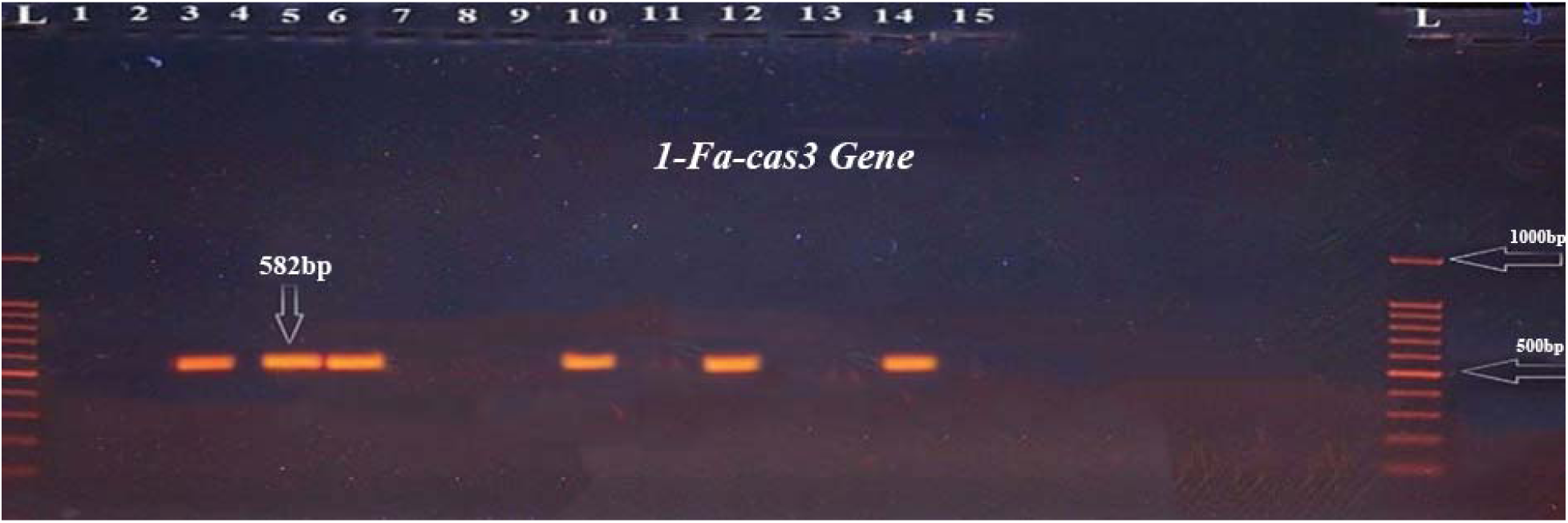
Gel electrophoresis of PCR product of *1-Fa-cas3*(582 bp) gene,L = ladder (100 -1500), melting temperature (Tm) of 55°C, 1% agarose gel, starting at 100 volts for 10 minutes and then reduced to 70 volts for 60 minutes, the (3,5,6,10,12,14 isolates represents of the positive results in samples for this gene.

**Fig (6):**
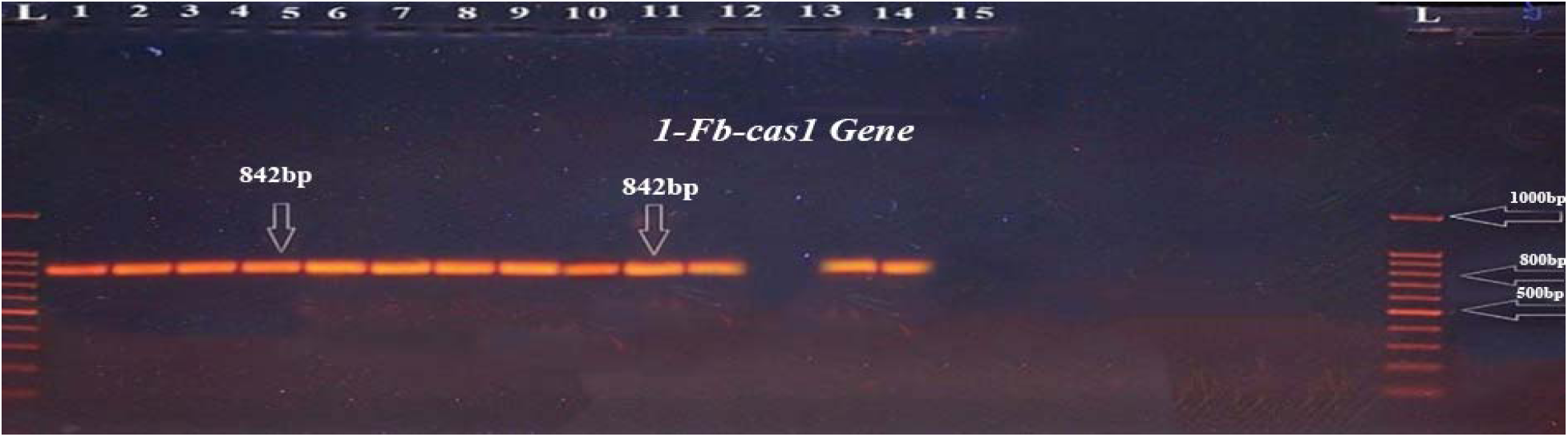
Gel electrophoresis of PCR product of *1-Fb-cas1*(842 bp) gene,L = ladder (100 -1500), melting temperature (Tm) of 55°C, 1% agarose gel, starting at 100 volts for 10 minutes and then reduced to 70 volts for 60 minutes, the (1-12,14 isolates represents of the positive results in samples for this gene.

**Fig (7):**
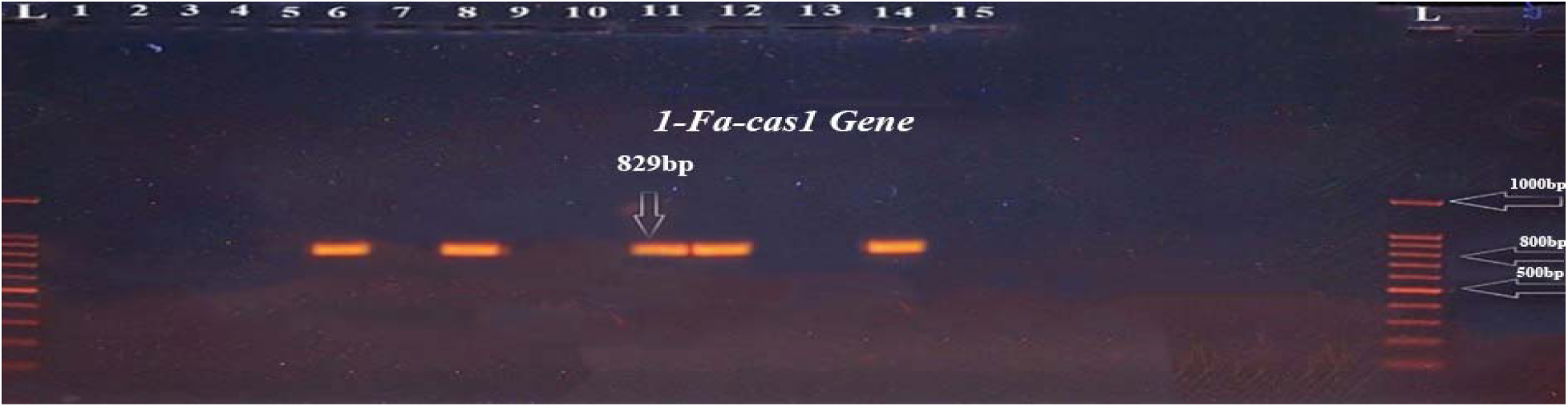
Gel electrophoresis of PCR product of *1-Fb-cas1*(829 bp) gene,L = ladder (100 -1500), melting temperature (Tm) of 55°C, 1% agarose gel, starting at 100 volts for 10 minutes and then reduced to 70 volts for 60 minutes, the (6,8,11,12,14 isolates represents of the positive results in samples for this gene.

#### Frequency of the CRISPR-Cas system among *Acinetobacter baumannii* isolates

In the present study, I-Fb-cas1 is the most commonly detected gene. I-Fa-cas1 and I-Fa-cas3 are less prevalent. The I-Fb subtype (cas1 and cas3) appears more active or retained in this isolates compared to the I-Fa subtype as in Fig(8).

**Fig. (8):**
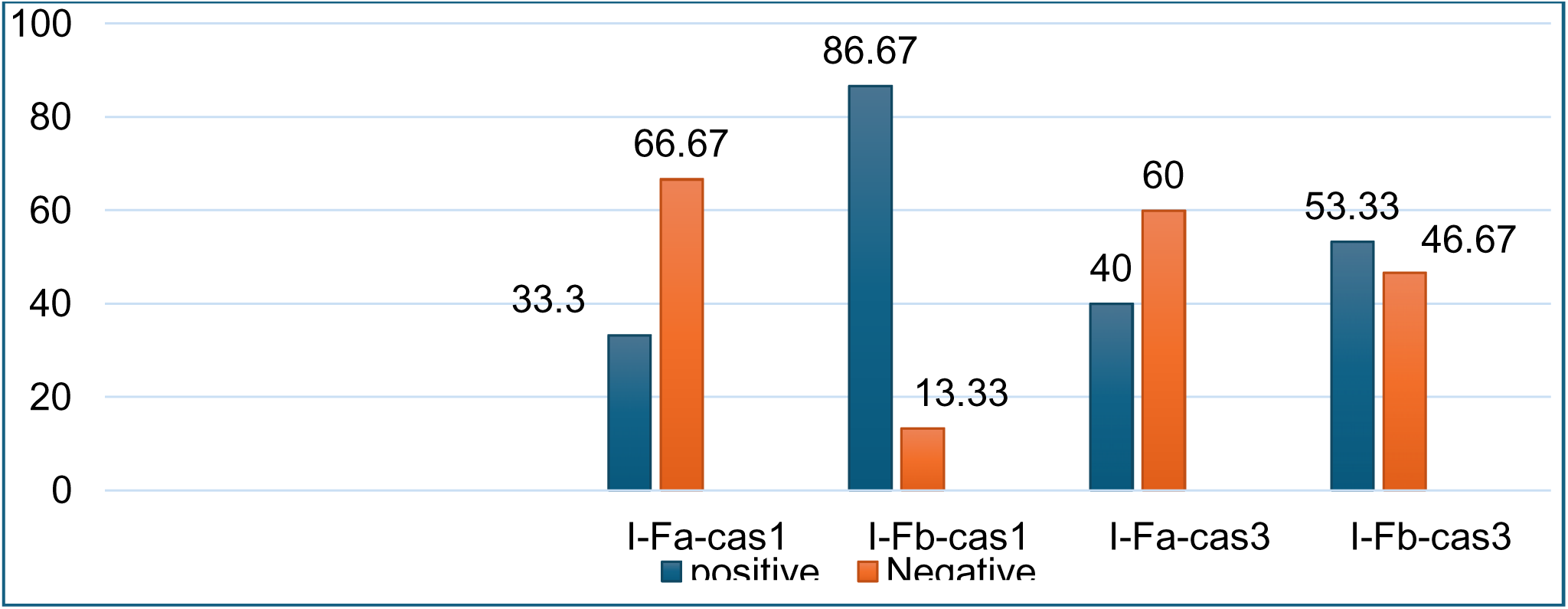
Frequency of CRISPR-Cas associated proteins among *Acinetobacter baumannii* isolates

#### Identification profile and Variability of CRISPR/Cas subtypes within *A. baumannii*

In the present study, the analysis of CRISPR/Cas systems in 15 clinical isolates of *Acinetobacter baumannii* revealed the presence of several CRISPR-associated genes, specifically I-Fa-cas1, I-Fb-cas1, I-Fa-cas3, and I-Fb-cas3, as detailed in Tables 8 and 9.The I-Fb-cas1 gene was the most frequently detected, present in 13 of the 15 isolates (86.7%). The I-Fa-cas1 gene was identified in 6 isolates (40%), while I-Fa-cas3 and I-Fb-cas3 were detected in 7 (46.7%) and 8 isolates (53.3%), respectively. Notably, two isolates (AB13 and AB15) lacked all tested CRISPR/Cas genes, whereas isolates AB12 and AB14 harboured the complete set of cas genes examined (I-Fa-cas1, I-Fb-cas1, I-Fa-cas3, and I-Fb-cas3). Several isolates, including AB1, AB2, AB4, AB7, and AB9, carried only the I-Fb subtype genes, particularly I-Fb-cas1 and, in some cases, I-Fb-cas3.Other isolates—such as AB3, AB5, AB6, AB8, and AB10—exhibited varied cas gene profiles, suggesting that horizontal gene transfer events may contribute to the diversity and distribution of CRISPR/Cas systems in *A. baumannii*. These findings highlight the genetic variability of CRISPR/Cas elements in this species and their potential involvement in genomic adaptation.

**Table (8):** Identification profile of CRISPR/Cas proteins in *Acinetobacter baumannii* isolates.

| No. of isolates | CRISPR/Cas proteins |  |  |  |
| --- | --- | --- | --- | --- |
|  | I-Fa-cas1 | I-Fb-cas1 | I-Fa-cas3 | I-Fb-cas3 |
| AB1 | - | + | - | + |
| AB2 | - | + | - | + |
| AB3 | - | + | + | - |
| AB4 | - | + | - | - |
| AB5 | - | + | + | + |
| AB6 | + | + | + | - |
| AB7 | - | + | - | + |
| AB8 | + | + | - | + |
| AB9 | - | + | - | - |
| AB10 | - | + | + | + |
| AB11 | + | + | - | - |
| AB12 | + | + | + | + |
| AB13 | - | - | - | - |
| AB14 | + | + | + | + |
| AB15 | - | - | - | - |

**Table (9):**
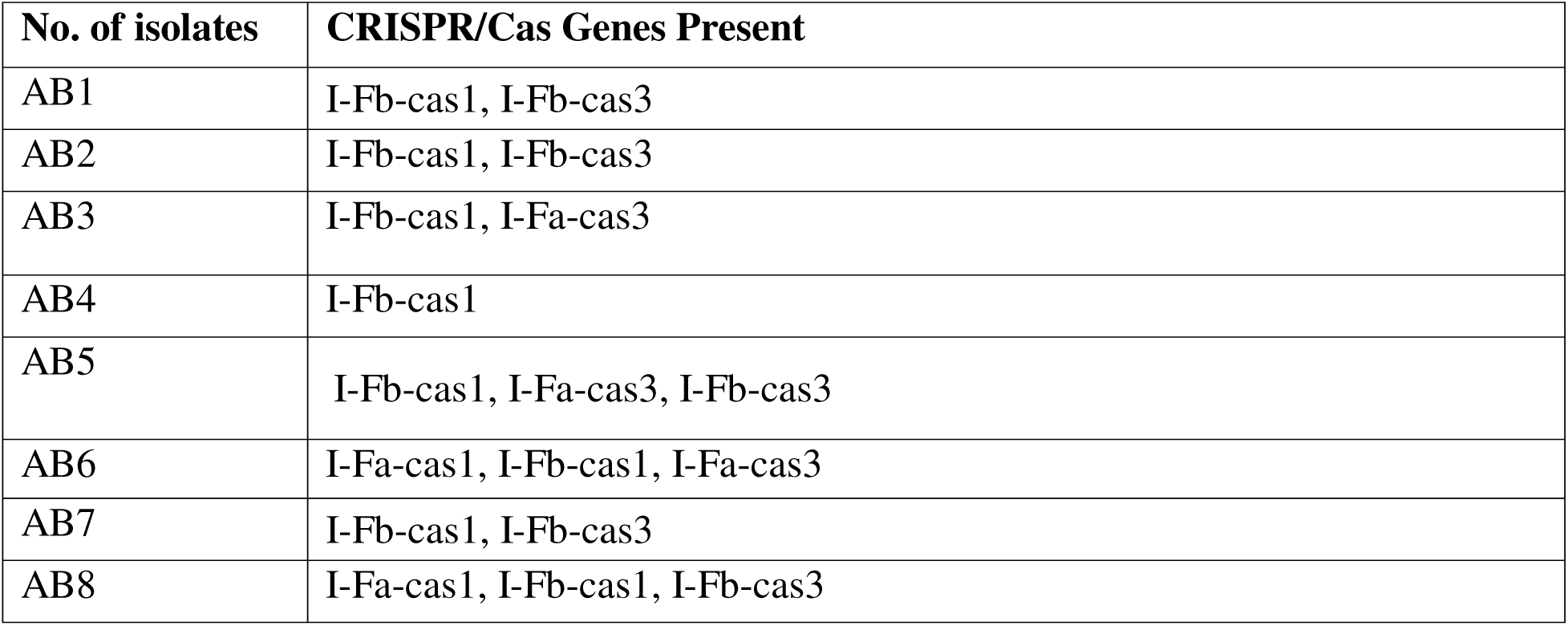
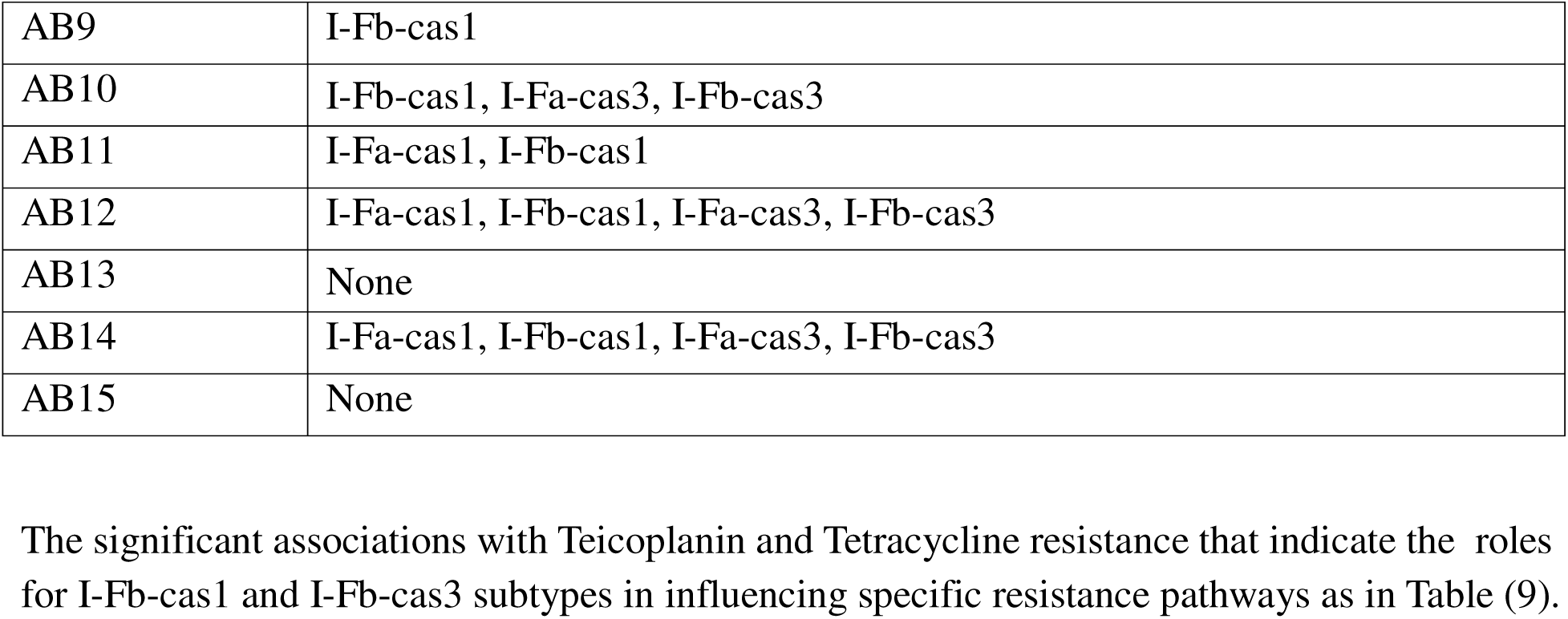
Distribution of CRISPR/Cas Genes Among Clinical Isolates of *Acinetobacter baumannii*.

#### Association Between Antimicrobial Susceptibility Profiles and CRISPR/Cas System Types in *Acinetobacter baumannii* Isolates

In the present study, the presence of different CRISPR-Cas subtypes—I-Fa-cas1, I-Fb-cas1, I-Fa-cas3, and I-Fb-cas3—was analyzed in relation to antimicrobial susceptibility profiles among 15 *Acinetobacter baumannii* clinical isolates. Statistical analysis was performed using p-values, with significance defined as *p* < 0.05.The I-Fa-cas1 gene was identified in a small subset of isolates (n = 5). All of these isolates exhibited resistance to multiple antibiotics, including benzylpenicillin, oxacillin, trimethoprim/sulfamethoxazole, and tigecycline. Partial sensitivity was observed for gentamicin, ciprofloxacin, and linezolid. However, no statistically significant association was found between the presence of I-Fa-cas1 and antibiotic susceptibility (*p* > 0.05).The I-Fb-cas1 gene was the most prevalent, detected in 13 isolates. These isolates demonstrated variable susceptibility across a range of antibiotics. Notably, some sensitivity was observed to clindamycin, linezolid, teicoplanin, and tigecycline. A statistically significant association was identified between the presence of I-Fb-cas1 and susceptibility to teicoplanin (*p* = 0.032).The I-Fa-cas3 gene was present in 6 isolates, most of which displayed high levels of resistance to the tested antibiotics. Intermediate responses were noted in a few isolates, particularly for ciprofloxacin and tigecycline. Nonetheless, the statistical analysis did not reveal any significant associations (*p* > 0.05).The I-Fb-cas3 gene was detected in 8 isolates and was associated with slightly higher susceptibility rates to tigecycline, linezolid, and clindamycin. A significant correlation was found between the presence of I-Fb-cas3 and increased susceptibility to tigecycline (*p* = 0.003), as shown in Table 10. These findings suggest that certain CRISPR-Cas subtypes may influence antimicrobial susceptibility patterns in *A. baumannii*, particularly in relation to tigecycline and teicoplanin.

**Table (10):**
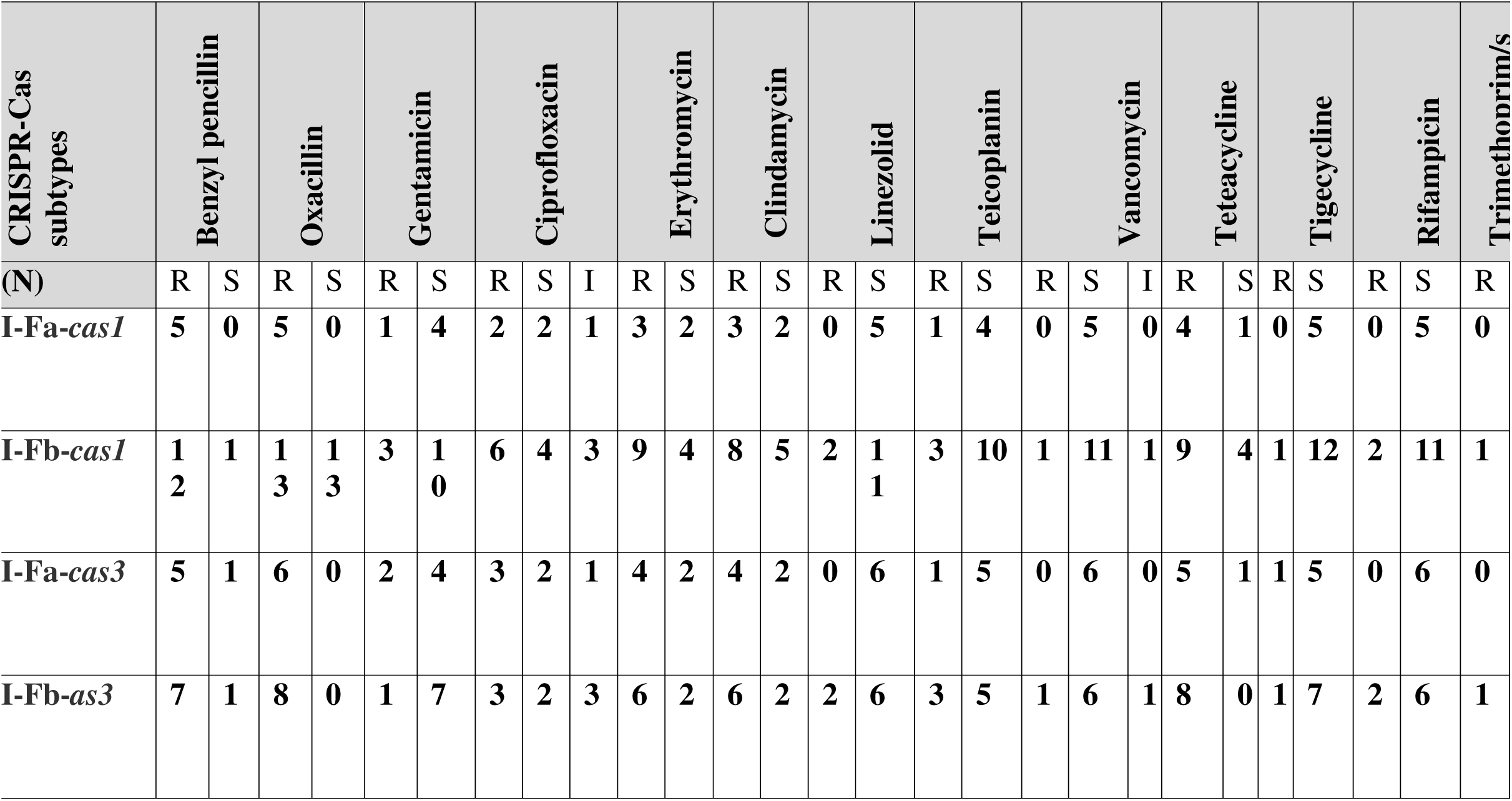
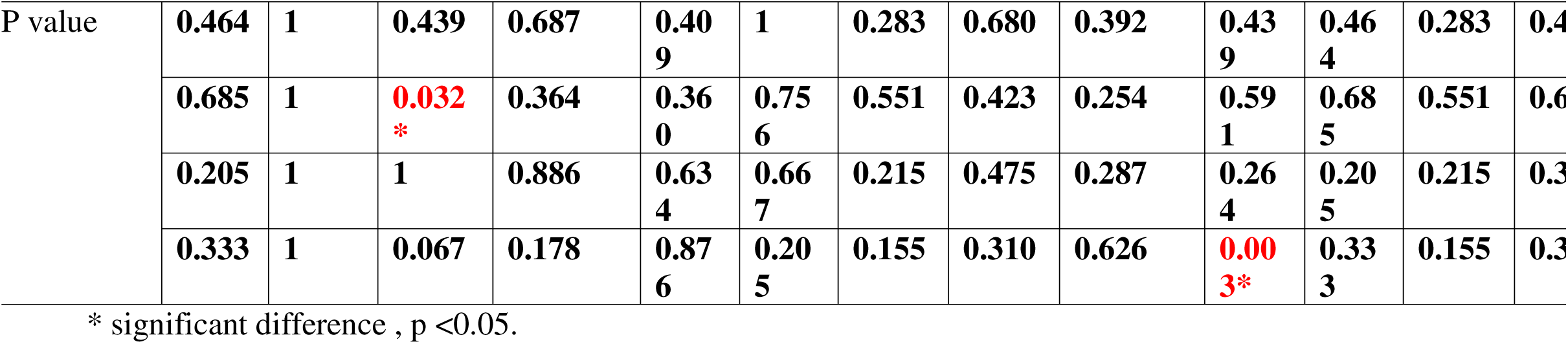
Correlation between antimicrobial susceptibility of *Acinetobacter baumannii* isolates and the presence of CRISPR/Cas Types.

#### Gene Expression Profile of *adeB*

In present study, Found in 10 out of 15 isolates, indicating that increase in adeB gene expression. While non-significant expression3 (20%)isolates. And downregulation 2(13.3%) isolates, indicating suppressed adeB expression as in Table(11).

**Table (11):**
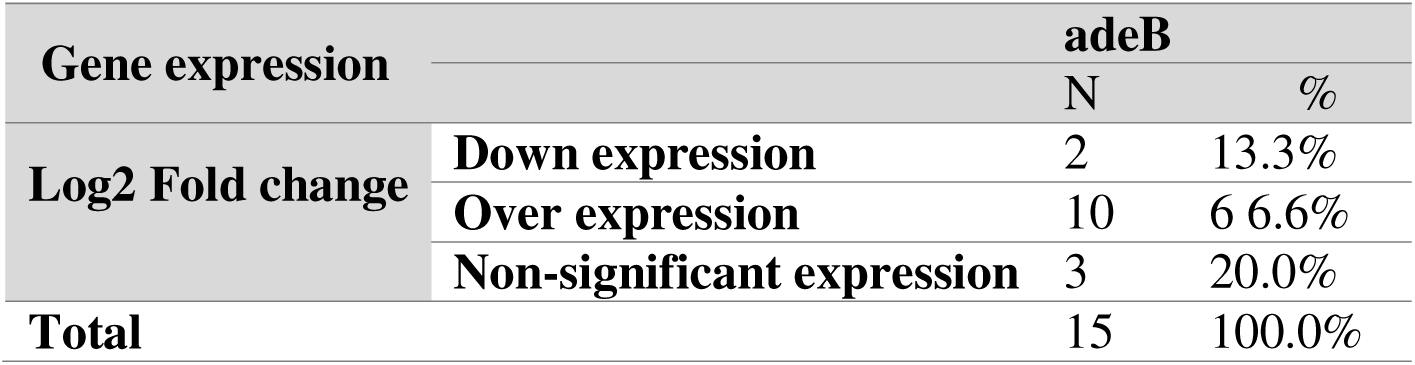
Distribution of adeB Efflux Pump Gene Expression Levels Among *Acinetobacter baumannii* Clinical Isolates.

#### 1.2.Correlation Between *adeB* Expression and CRISPR-Cas Gene Profiles in *A. baumannii*

In present study, the relationship between CRISPR-Cas gene profiles and the expression level of the adeB efflux pump gene in *Acinetobacter baumannii*(Downregulated adeB expression was observed in 2 isolates that carried all four CRISPR-Cas genes: *I-Fa-cas1*, *I-Fb-cas1*, *I-Fa-cas3*, and *I-Fb-cas3*. This indicates that the presence of a complete CRISPR-Cas system may play a regulatory role in suppressing the expression of the *adeB* efflux pump, while Overexpressed adeB expression was found in 10 isolates, which predominantly carried I-Fb-cas1 and I-Fb-cas3, but lacked theCRISPR-Cas gene set. that incomplete CRISPR-Cas systems and no regulatory effect on *adeB* expression, resulting in upregulation of this efflux pump **and** enhanced drug resistance as in table(12).

**Table (12):**
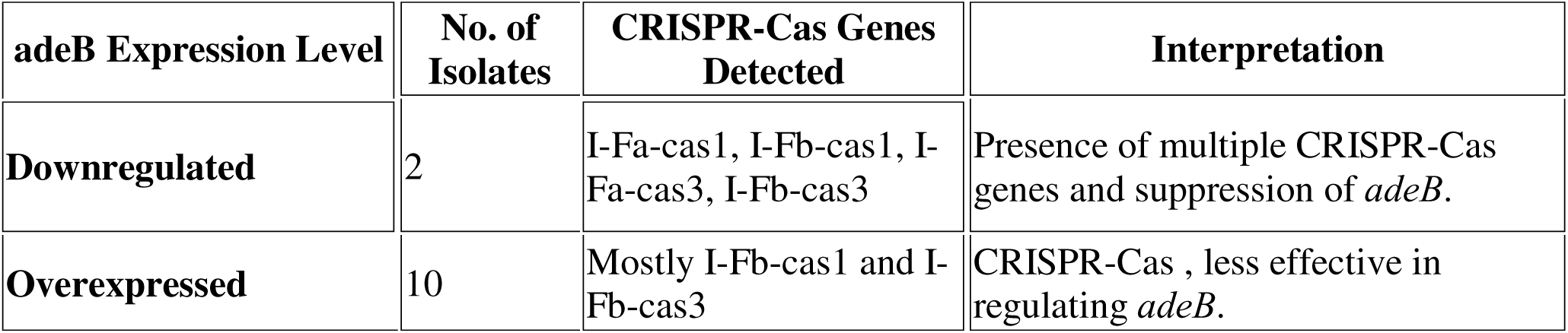
Association Between CRISPR-Cas Profiles and adeB Expression in *A. baumannii*.

In the present study, it was found that certain CRISPR-Cas configurations could influence antibiotic resistance mechanisms in *A. baumannii* via modulation of efflux pump genes as in Fig(9).showed that Downregulated adeB in 2 isolates, CRISPR-Cas profile present including(I-Fa-cas1, I-Fb-cas1, I-Fa-cas3, I-Fb-cas3), while overexpressed adeB in 10 isolates, CRISPR-Cas profile present in ( I-Fb-cas1 and I-Fb-cas3, Non-significant Expression in 3 isolates

**Figure (9):**
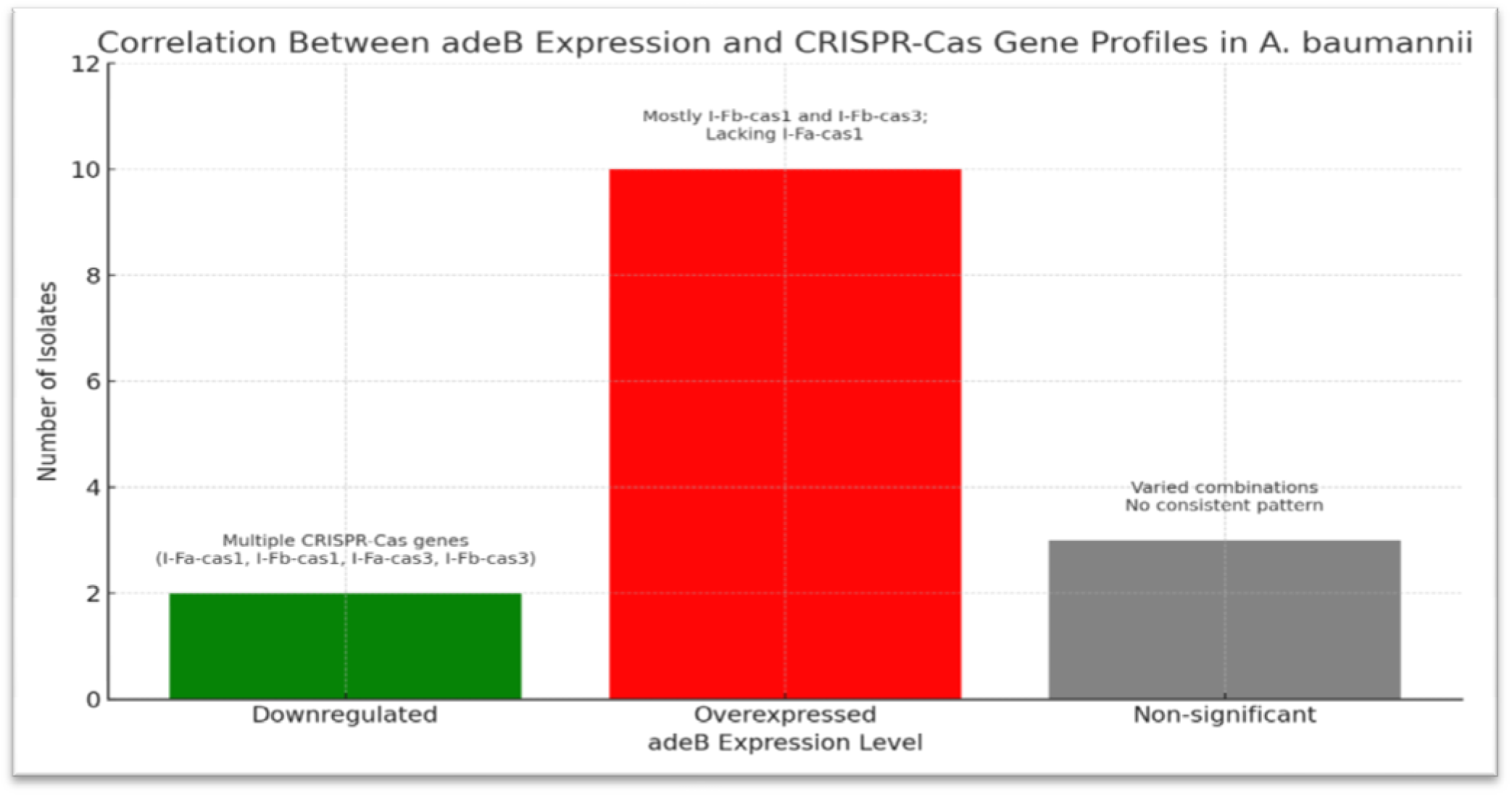
Correlation Between *adeB* Expression and CRISPR-Cas Gene Profiles in *A. baumannii*

## Discussion

In the present study, *Acinetobacter baumannii* was isolated from 15(15%) out of 100 clinical wound swab specimens as inFig(1). This isolation rate is consistent with previous reports indicating that *A. baumannii* is a common nosocomial pathogen, especially in wound infections and intensive care units (Sahiledengle *et al*., 2023; Tiwari *et al*., 2024). The identification of *A. baumannii* was confirmed by PCR amplification of the intrinsic *blaOXA-51* gene as in Fig(3), which was detected in 100% of isolates. This gene remains a reliable molecular marker for distinguishing *A. baumannii* from other Acinetobacter species (Antunes *et al*., 2022).The antimicrobial susceptibility testing (AST) of 15 *A. baumannii* isolates as in (Table 2,Fig 2) revealed high resistance rates to several commonly used antibiotics. Notably, oxacillin (100%), benzylpenicillin (93.3%), and erythromycin (73.3%) exhibited the highest levels of resistance. These findings are in agreement with recent data showing widespread resistance of *A. baumannii* to β-lactams and macrolides (Chen *et al*., 2023). Resistance to ciprofloxacin (53.3%) and clindamycin (60%) further underscores the challenge in treating *A. baumannii* infections using fluoroquinolones or lincosamides, especially in multidrug-resistant (MDR) strains. and a high percentage of isolates were sensitive to tigecycline (93.3%), trimethoprim/sulfamethoxazole (93.3%), and rifampicin (86.7%), consistent with recent studies suggesting these agents may still retain activity against resistant strains (Yousefi *et al*., 2024; Mohamed *et al*., 2023). linezolid (86.7%), teicoplanin (73.7%), and vancomycin (80%) showed promising sensitivity profiles, although vancomycin is traditionally considered less effective against Gram-negative organisms due to poor outer membrane penetration (Pereira *et al*., 2024). However, its intermediate susceptibility rate (13.3%) in this study the variable permeability or non-classical mechanisms of action. The statistical significance observed for teicoplanin, tigecycline, and rifampicin (p < 0.05) supports their potential therapeutic role, although resistance to tetracycline (66.7%) highlights the continued evolution of resistance even to older agents. The presence of CRISPR-Cas system components was analyzed among the 15 *A. baumannii* isolates to explore their potential role in regulating resistance mechanisms. All isolates were positive for *blaOXA-51*, affirming species identity. The I-Fb-cas1 gene was detected in 86.7% of isolates, showing a significant association (p = 0.005), while I-Fa-cas1 was found in 33.3%, indicating strain variability or environmental influence. I-Fb-cas3 and I-Fa**-**cas3 were present in 53.3% and 40% of isolates, respectively.These findings align with recent studies that CRISPR-Cas systems in *A. baumannii* may be variably expressed, potentially affecting their ability to interfere with foreign DNA or modulate endogenous genes such as efflux pumps (Rathinam *et al*., 2022; Martens *et al*., 2023).The observed co-presence of multiple CRISPR-Cas components in some isolates. The results showed that isolates with complete CRISPR-Cas profiles (I-Fa-cas1, I-Fb-cas1, I-Fa-cas3, I-Fb-cas3) were associated with downregulation of the *adeB* efflux gene, that suppressive effect. The isolates with only I-Fb elements exhibited overexpression of *adeB*, indicating that partial CRISPR-Cas systems may be insufficient to regulate efflux mechanisms. The CRISPR-Cas elements may contribute to the modulation of antimicrobial resistance genes, as highlighted in recent molecular investigations (Gholizadeh *et al*., 2023).The significant associations between CRISPR-Cas profiles and antibiotic resistance, particularly in relation to teicoplanin and tetracycline, the interference with horizontal gene transfer or direct interaction with resistance gene promoters. The current study sought to investigate the relationship between the presence of CRISPR-Cas subtypes and antimicrobial resistance patterns among *Acinetobacter baumannii* isolates. Our findings indicate variable associations between specific CRISPR-Cas elements and susceptibility to individual antibiotics, with certain statistically significant correlations, particularly in relation to teicoplanin and tetracycline resistance.Among the CRISPR-Cas subtypes, I-Fb-cas3 showed a significant association with tetracycline **resistance** (*p* = 0.003), that indicate the role in modulating resistance pathways specific to tetracycline-class antibiotics. This aligns with the findings of Gholizadeh *et al*. (2023), who reported subtype-specific CRISPR-Cas elements potentially influencing horizontal gene transfer and the expression of antimicrobial resistance genes. Similarly, a significant association was observed between I-Fa-cas3 and teicoplanin susceptibility (*p* = 0.032), hinting at a possible protective or regulatory function of this subtype in reducing glycopeptide resistance.While no statistically significant correlations were found between other CRISPR-Cas types (I-Fa-cas1 or I-Fb-cas1) and resistance to β-lactams (benzylpenicillin, oxacillin), aminoglycosides (gentamicin), fluoroquinolones (ciprofloxacin), or macrolides (erythromycin). These results indicate that the role of CRISPR-Cas systems in resistance modulation may be gene- and antibiotic-specific, rather than broad-spectrum.These observations are consistent with recent studies by Rathinam *et al*. (2022) and Martens *et al*. (2023), which propose that incomplete or variably expressed CRISPR-Cas systems in *A. baumannii* might have limited functionality in regulating the full range of resistance determinants but may still play roles in specific regulatory circuits.the functional role of CRISPR-Cas elements in antibiotic resistance, the expression levels of the ***adeB*** efflux pump gene were analyzed. Of the 15 *A. baumannii* isolates, 10 isolates (66.6%) showed overexpression of adeB, consistent with high efflux activity and potential multidrug resistance. Only 2 isolates (13.3%) exhibited downregulated expression, while 3 (20%) had no significant change. The two isolates with *adeB* downregulation carried a complete set of CRISPR-Cas genes: I-Fa-cas1, I-Fb-cas1, I-Fa-cas3, and I-Fb-cas3. This strongly suggests that when fully present and potentially functional, the CRISPR-Cas system may act as a negative regulator of the efflux pump system. This regulatory capacity may occur through direct targeting of resistance-associated mRNA transcripts or interference with gene promoters, as hypothesized in prior molecular studies (Makarova *et al*., 2021; Jiang & Doudna, 2022).The isolates with overexpressed *adeB* harbored primarily I-Fb-associated CRISPR-Cas genes, which appear to lack the regulatory robustness of the complete subtype combination. These findings support the emerging concept that incomplete or partial CRISPR-Cas arrays may be ineffective in regulating resistance genes, thereby allowing unregulated expression of efflux systems like *adeB.*(Karvelis *et al*., 2023; Zarei *et al*., 2024).

## Conclusion

In this study, CRISPR-Cas gene profiles may play a suppressive role in regulating adeB efflux gene expression, thereby influencing the resistance phenotype of *A. baumannii.* These findings highlight the dual role of CRISPR-Cas systems in both genetic defence and antimicrobial resistance modulation, suggesting their potential as targets for diagnostic and therapeutic development. The interplay between CRISPR-Cas elements and efflux pump regulation represents a novel and promising area of antimicrobial resistance research. Targeting these regulatory systems may open new therapeutic strategies to combat multidrug-resistant A. baumannii, as highlighted by newer approaches using CRISPR-based antimicrobials.

## Data Availability

All data produced in the present work are contained in the manuscript

## Acknowledgement

I would like to thank the staff (Al-Hilla General Teaching Hospital for helping).

## Financial support and sponsorship

Nil.

## Conflict of interest

NO.

## Notes

### Competing Interest Statement

The authors have declared no competing interest.

### Funding Statement

not receive any funding

### Author Declarations

Ethical approval for this study was obtained from the ethical committee at Hilla Surgical Teaching Hospital. This study was also approved by a local ethics committee at the College of Medicine, University of Babylon. and the hospital ethics committee under document number [IRB: 399-4/4/2025].

